# Estimating white matter hyperintensities volume in individuals with stroke using T1-weighted images

**DOI:** 10.1101/2025.10.22.25338564

**Authors:** Mahir H. Khan, Stuti Chakraborty, Jennifer K. Ferris, Lara A. Boyd, Mohamed Salah Khlif, Amy Brodtmann, Michael R. Borich, James H. Cole, Steven C. Cramer, Niko H. Fullmer, Jeanette R. Gumarang, Hosung Kim, Amisha Kumar, Octavio Marin-Pardo, Susan M. Murphy, Emily R. Rosario, Heidi M. Schambra, Grace C. Song, Sook-Lei Liew

## Abstract

Stroke recovery outcomes vary across individuals, motivating the search for biomarkers that can improve prediction. White matter hyperintensities (WMH) volume is a leading biomarker candidate, with FLAIR MRI typically used for WMH segmentation; however, T1-weighted (T1) MRI is often more available. Therefore, we evaluated the performance of two automated WMH segmentation methods (WMH-SynthSeg and SAMSEG) to determine whether WMH volume can be reliably estimated using T1 alone. We analyzed imaging data from 227 stroke patients across three datasets spanning early subacute to chronic recovery, each with gold-standard WMH masks and stroke lesion masks manually traced on available T1 and FLAIR scans. WMH was segmented using T1 only as input to WMH-SynthSeg and SAMSEG, as well as using both T1 and FLAIR as input to SAMSEG, as previously implemented in stroke recovery research. Automated WMH segmentations were compared to the gold-standard WMH mask: accuracy was assessed using Dice similarity index (SI) and cluster-level false negative ratio, while agreement was assessed using intraclass correlation, Pearson’s correlation, and volume ratio. We used linear mixed-effects models to evaluate whether SI was influenced by factors such as WMH volume, stroke lesion volume, WMH contrast, age, sex, and days since stroke, with dataset as a random effect. WMH-SynthSeg using T1-only input produced more accurate and reliable WMH segmentations compared to SAMSEG with T1-only input and performed comparably to SAMSEG using both T1 and FLAIR input. WMH-SynthSeg using T1-only input may be used for WMH volume estimation in stroke recovery research in the absence of multimodal imaging.

**Graphical Abstract:** 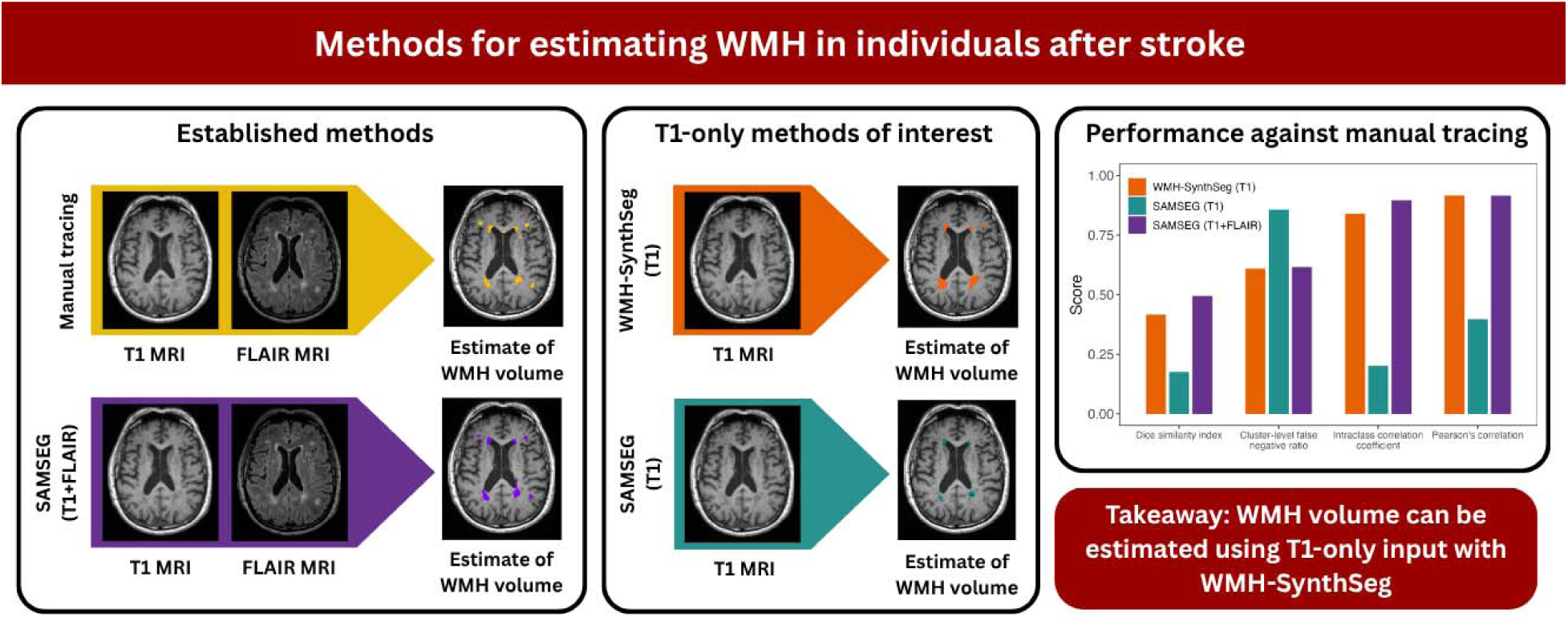

**Highlights:**

– WMH volume, often assessed via multimodal imaging, predicts post-stroke outcomes
– T1-only methods would facilitate WMH analysis if multimodal MRI is unavailable
– It is unclear how T1-only methods perform in brains with stroke lesions
– We show T1-based WMH-SynthSeg estimates strongly agree with gold standard methods
– Accuracy was stable across stroke lesion sizes but varied with WMH volume/contrast

## 1. Introduction

Stroke recovery outcomes differ greatly between individuals and are influenced by factors such as volume and location of focal stroke damage (Martin et al., 2024; Sperber et al., 2023). This variability presents significant challenges for predicting recovery (Bonkhoff and Grefkes, 2021), which is important for treatment planning and helping patients and their families adapt. To this end, there is growing interest in identifying biomarkers that can improve outcome prediction and guide more targeted rehabilitation approaches (Boyd et al., 2017).

Predictive biomarkers can be based on the acute infarct but can also be derived from measures of whole brain health following the stroke. One imaging biomarker of interest is white matter hyperintensities (WMH) of presumed vascular origin, a neuroimaging finding that reflects both age-related and disease-based vascular injury and small vessel disease (Brown et al., 2021; Duering et al., 2023). These WMHs appear as areas of abnormal signal intensity within white matter, best seen on MRI, primarily because these regions typically exhibit higher water content and interstitial fluid mobility before more permanent changes such as demyelination occur (Wardlaw et al., 2015). In the context of stroke recovery, higher WMH volume has been associated with poorer functional outcomes after stroke, independent of biomarkers related to focal stroke damage (Bonkhoff et al., 2022; Ferris et al., 2024; Hong et al., 2021), and with poorer response to an approved stroke recovery therapy (Schwarz et al., 2025).

Recent advances in computational modeling have enabled the development of sophisticated algorithms capable of automatically segmenting WMH across different MRI modalities. These approaches include cross-modality and multi-modality training frameworks that leverage information from multiple imaging sequences to improve accuracy and generalizability. Notably, two promising methods in this domain are SAMSEG (Sequence Adaptive Multimodal SEGmentation) and WMH-SynthSeg (Cerri et al., 2021; Laso et al., 2024). Both are freely available neuroimaging algorithms designed for robust, automated WMH segmentation and offer flexibility with input modalities, supporting images such as fluid-attenuated inversion recovery (FLAIR) and T1-weighted (T1) MRI. Each was trained and validated on large and diverse datasets, demonstrating robust performance across heterogeneous clinical populations (Johnson et al., 2025; Kern et al., 2022; Kim et al., 2025; Malla et al., 2025; Ortega-Cruz et al., 2025), however these methods have not been widely studied in stroke where varying stroke lesions may affect segmentation accuracy.

Automated WMH segmentation pipelines are particularly valuable for large-scale collaborative efforts in stroke recovery research, such as the ENIGMA Stroke Recovery Working Group, which harnesses large, harmonized datasets to refine prognostic models and validate biomarkers like WMH volume (Ferris et al., 2024; Liew et al., 2022b). FLAIR MRI sequences are particularly effective at capturing WMH, as they suppress the signal from cerebrospinal fluid and thereby accentuate abnormalities in white matter water content (Wardlaw et al., 2015). However, WMHs are also visible on T1-weighted MRI, where they appear hypointense and are thought to reflect more advanced tissue degeneration (Riphagen et al., 2018). Prior studies have demonstrated that despite these modality-specific differences, WMH volumes derived from FLAIR and T1 are highly correlated (Wei et al., 2019). Importantly, for multi-site stroke studies such as ENIGMA, where data from different studies is harmonized for greater statistical power and generalizability, T1 MRI is more widely available and reliable across sites, partly due to greater standardization across sites and broader relevance across acute to chronic recovery stages (Liew et al., 2022a). The ability to accurately estimate WMH from T1 images alone could therefore broaden participant inclusion, increase statistical power, and facilitate further investigation into the role of WMH volume in stroke recovery.

In the present study, we aimed to compare WMH volume estimation in stroke survivors using only T1 MRI with WMH-SynthSeg and SAMSEG. We further compared the quality of these T1-only segmentations against a reference method of using both T1 and FLAIR with SAMSEG, previously shown to be reliable (Ferris et al., 2023). Demonstrating reliable WMH segmentation from T1 alone could increase the number of participants eligible for WMH-related studies for stroke, particularly in large-scale or retrospective datasets, when FLAIR is unavailable.

## 2. Methods

### 2.1. Participants

Neuroimaging and demographic data were pooled from 227 individuals across three independent datasets. All relevant imaging parameters can be found in Supplementary Table 1.

The first dataset (Dataset 1) comprises a chronic (greater than 6 months since stroke) stroke cohort of 43 individuals collected at the Brain Behavior Laboratory of the University of British Columbia (UBC), as described previously (Ferris et al., 2023). MRI was collected using a 3T Phillips Achieva or Elition scanner. A T1 3D magnetization-prepared rapid gradient-echo (MPRAGE) scan and a FLAIR scan were acquired.

The second dataset (Dataset 2) represents a late subacute (3 months since stroke) stroke cohort of 120 participants collected from the Cognition and Neocortical Volume after Stroke (CANVAS) study. Inclusion criteria have been previously published (Brodtmann et al., 2014; Ferris et al., 2023). T1 MPRAGE and FLAIR scans were collected using a 3T Siemens Tim Trio scanner.

The final dataset (Dataset 3) constituted an early subacute (21 days since stroke) cohort of 64 individuals collected from Casa Colina Hospital, Emory University, and NYU Langone Health as part of the Global Brain Health After Stroke study. Study participants were recruited according to the following inclusion criteria: (1) at least one major upper extremity muscle group with a manual muscle test score of less than 5/5, (2) written and oral proficiency in English or Spanish, and (3) willingness to complete study procedures. Exclusion criteria included traumatic brain injury or presence of major musculoskeletal or secondary neurological condition. T1 MPRAGE and FLAIR scans were collected using 3T Siemens MAGNATOM Verio and Siemens MAGNATOM PrismaFit scanners.

Ethical approval was granted by the appropriate institutional review boards for each dataset: University of British Columbia Clinical Research Ethics Board (Dataset 1); Austin Hospital Research Ethics Committee, Box Hill Hospital Research Ethics Committee, and Royal Melbourne Hospital Research Ethics Committee (Dataset 2); and the University of Southern California Health Sciences Campus Institutional Review Board and the local ethics board for Casa Colina Hospital, Emory University, and New York University Langone Health (Dataset 3). Written informed consent was obtained from all participants in each study.

### 2.2. WMH and stroke lesion estimation

“Gold standard” WMH masks were generated for each dataset. In Dataset 1, masks were generated using SABRE, a semi-automated brain segmentation tool that leverages multimodal imaging, followed by manual correction (Ferris et al., 2023; Ramirez et al., 2011). In Datasets 2 and 3, masks were initially generated using SAMSEG, an unsupervised method that performs image segmentation using parametric Bayesian modeling on single and multi-contrast MRI data without the need for any preprocessing (Cerri et al., 2021), followed by manual correction. Separately, stroke lesions were traced manually in all datasets.

### 2.3. Automated WMH segmentation

We used two fully automated WMH segmentation pipelines implemented in FreeSurfer v8.0.0: WMH-SynthSeg (Laso et al., 2024) and SAMSEG (Cerri et al., 2021) using only T1 images for comparison. WMH-SynthSeg is an unsupervised segmentation method that uses a previously-trained convolutional neural network architecture to label different tissue classes from brain MRI scans of any contrast and resolution without the need for retraining, re-tuning, or preprocessing (Laso et al., 2024).

We ran the GPU-accelerated version of WMH-SynthSeg using only T1 images. WMH masks were generated by binarizing the voxels labeled as WMH in the resulting brain tissue segmentation volume; these will be referred to as *WMH-SynthSeg (T1) WMH masks*. SAMSEG was also run with only T1 images as input, using previously recommended parameters (Ferris et al., 2023), including the white matter lesion extension and a lesion threshold of 0.1; yielding WMH masks are referred to as *SAMSEG (T1) WMH masks*. Additionally, the WMH-SynthSeg (T1) and SAMSEG (T1) WMH masks were corrected to exclude overlapping voxels with the stroke lesion masks traced a-priori.

As a reference point, we also generated WMH masks using SAMSEG as described in Ferris et al. (2023), utilizing both T1 and FLAIR images as input, the white matter lesion extension, a lesion threshold of 0.1, and correction using stroke lesion masks (Ferris et al., 2023). This protocol was found to be a reliable method for estimating WMH in people with stroke, and was later validated when associating WMH volume to motor outcomes after stroke (Ferris et al., 2024). WMH masks obtained using this method are hereby referred to as *SAMSEG (T1+FLAIR) WMH masks*.

A schematic summarizing how each WMH mask was produced is shown in Figure 1. For thoroughness, we also conducted an exploratory analysis to evaluate WMH segmentation when FLAIR was used as the sole input. Methods and results can be found in the *Supplementary Materials*.

**Figure 1:**
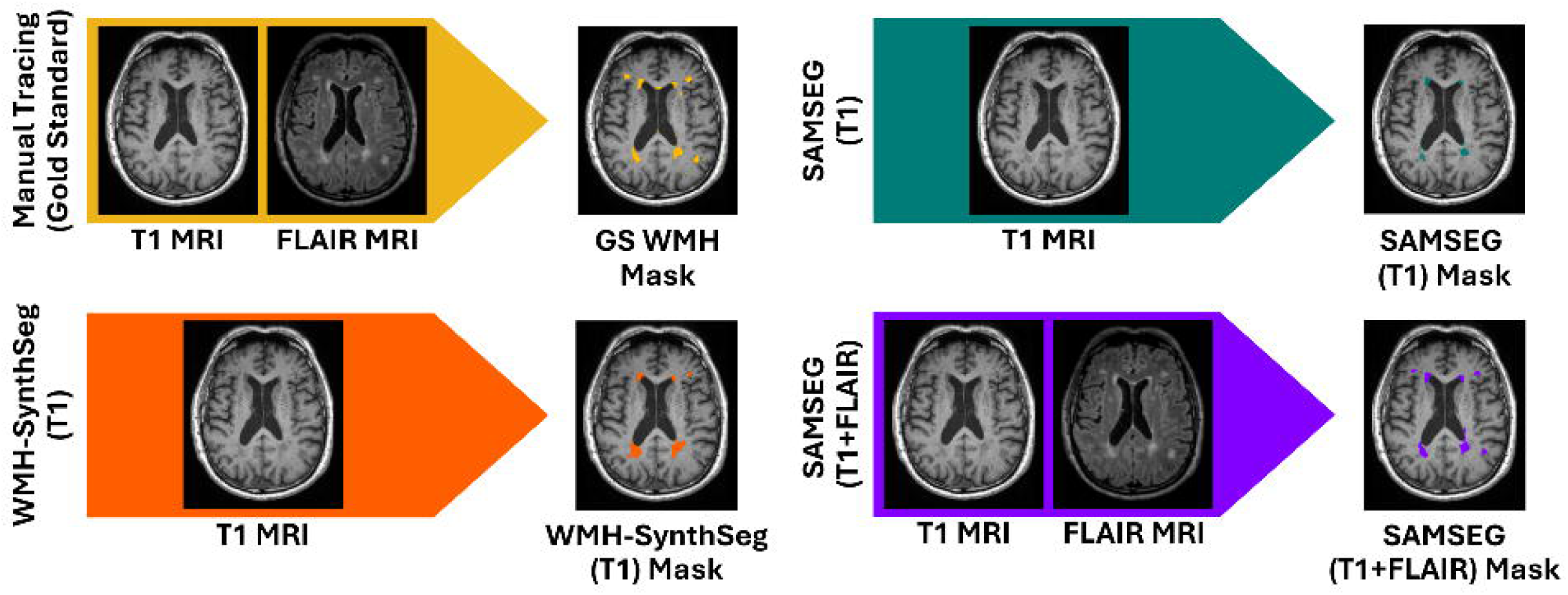
Schematic demonstrating the algorithm and input combinations used to produce WMH masks. The gold standard WMH masks were produced using semi-automated approaches followed by manual tracing on T1 and FLAIR images. WMH-SynthSeg (T1) and SAMSEG (T1) masks were produced using T1-only input to WMH-SynthSeg and to SAMSEG, respectively. SAMSEG (T1+FLAIR) masks were produced using T1 and FLAIR inputs to SAMSEG.

### 2.4. WMH contrast quantification

We conducted an exploratory analysis to examine whether the T1 intensity contrast between WMH tissue and remaining white matter (WM) tissue influenced automated segmentation performance. For each subject, we generated WM masks by binarizing voxels labeled as cerebral WM in the T1-only WMH-SynthSeg segmentation output. We then modified this mask by removing voxels that overlapped with the gold-standard WMH and stroke lesion masks. We obtained the median T1 intensities of the WMH and WM masks. We then calculated the WMH-WM T1 intensity ratio as the median T1 intensity of the WMH mask normalized to the median T1 intensity in the WM mask. Lower ratios reflect greater differentiation between WMH and WM, which we hypothesized would be associated with improved segmentation accuracy.

### 2.5. Comparison of segmentation methods

We compared segmentation quality for each automated segmentation by computing metrics including Dice similarity index (SI), cluster-level false negative ratio (FNRc), intraclass correlation (ICC), Pearson’s correlation, and volume ratios against the gold standard WMH mask. The SI quantifies voxel-wise overlap between masks, where higher SI indicates greater degree of overlap. The FNRc captures differences in cluster-level overlap, and higher FNRc values indicate more missed or fragmented lesion clusters (Griffanti et al., 2016). The ICC and Pearson’s correlation assess the agreement in total WMH volume between each algorithm and the gold standard. The ICC scores, calculated using the *psych* package in R, were interpreted using the following scale: poor reliability = less than 0.5, moderate reliability = 0.5 to 0.75, good reliability = 0.75 to 0.9, and excellent reliability = greater than 0.9 (Koo and Li, 2016). Lastly, volume ratio was calculated as estimated WMH volume divided by the gold standard WMH volume, providing an estimate of over- or under-segmentation by each algorithm.

We tested for differences in volume ratio between segmentation methods. Volume ratios were non-normally distributed and therefore were assessed using the Kruskal–Wallis test, a non-parametric alternative to one-way ANOVA, via the *sjstats* package in R. Pairwise comparisons were subsequently performed using Wilcoxon rank-sum tests with Bonferroni correction to adjust for multiple comparisons.

We also explored whether WMH-WM intensity ratio influenced segmentation quality. First, we assessed differences in WMH-WM ratios between datasets using the Kruskal-Wallis test due to non-normal distributions in each dataset, then used the Wilcoxon rank-sum test to do pairwise comparisons. We then used standardized linear mixed-effects regression modeling to test whether WMH-WM intensity ratio was related to age, sex, days since stroke, gold-standard WMH volume, or stroke lesion volume, while including a random effect for each dataset. Dataset and days since stroke were found to be highly collinear (variance inflation factor [VIF] > 5) as each dataset was collected at a different recovery phase. We thus retained days since stroke as a fixed effect and modeled dataset as a random intercept to account for unmeasured differences across datasets, such as variations in scanners, imaging protocols, or other dataset-specific factors that may not have been directly measured. The WMH-WM intensity ratio, days since stroke, WMH volume, and stroke lesion volume were log-transformed to address skewed distributions and improve model assumptions of linearity and normality. Subsequently, model diagnostics confirmed that assumptions of linearity, normality of residuals, and appropriate specification of random effects were met.

Lastly, we used standardized linear mixed-effects regression to test whether segmentation quality was related to WMH volume, stroke lesion volume, and WMH-WM intensity ratio, controlling for age, sex, and days since stroke. We again included a random effect of dataset to account for differences between datasets. Log transformations were again used, and diagnostic checks confirmed that model assumptions were not violated.

## 3. Results

The study included 227 participants from three datasets. Dataset 1 consisted of 43 subjects (30.2% female) with a mean (± standard deviation (SD)) age of 65.3 ± 8.8 years at baseline visit, assessed on average 2078.3 ± 1768.8 days since stroke. Dataset 2 included 120 individuals (32% female), who had a mean age of 67.7 ± 11.6 years, and evaluated at an average of 99.0 ± 23.1 days since stroke. Dataset 3 comprised 64 participants (54% female), who had a mean age of 58.0 ± 13.7 years at baseline visit, and assessed on average at 24.0 ± 8.9 days since stroke.

### 3.1. WMH-SynthSeg (T1) performs comparably to SAMSEG (T1 + FLAIR)

We compared the performance of WMH-SynthSeg (T1) and SAMSEG (T1) with SAMSEG (T1+FLAIR) against the gold standard WMH masks across all three datasets (Fig. 2). WMH-SynthSeg (T1) achieved moderate to good performance across all datasets in relation to the gold standard (SI = 0.42 ± 0.20, FNRc = 0.61 ± 0.19, ICC = 0.84, *r* = 0.92). SAMSEG (T1) had poor performance across the board (SI = 0.18 ± 0.20, FNRc = 0.86 ± 0.18, ICC = 0.20, *r* = 0.40). Meanwhile, SAMSEG (T1+FLAIR) also achieved good performance across all datasets in relation to the gold standard (SI = 0.50 ± 0.26, FNRc = 0.62 ± 0.25, ICC = 0.90, r = 0.92). WMH-SynthSeg (T1) showed the most consistent performance in individual cohorts; detailed results are provided in *Supplementary Materials*. Heatmaps of the segmentations, including overlapping voxels with the gold standard, as well as false positives and false negatives, are illustrated in Figure 3.

**Figure 2:**
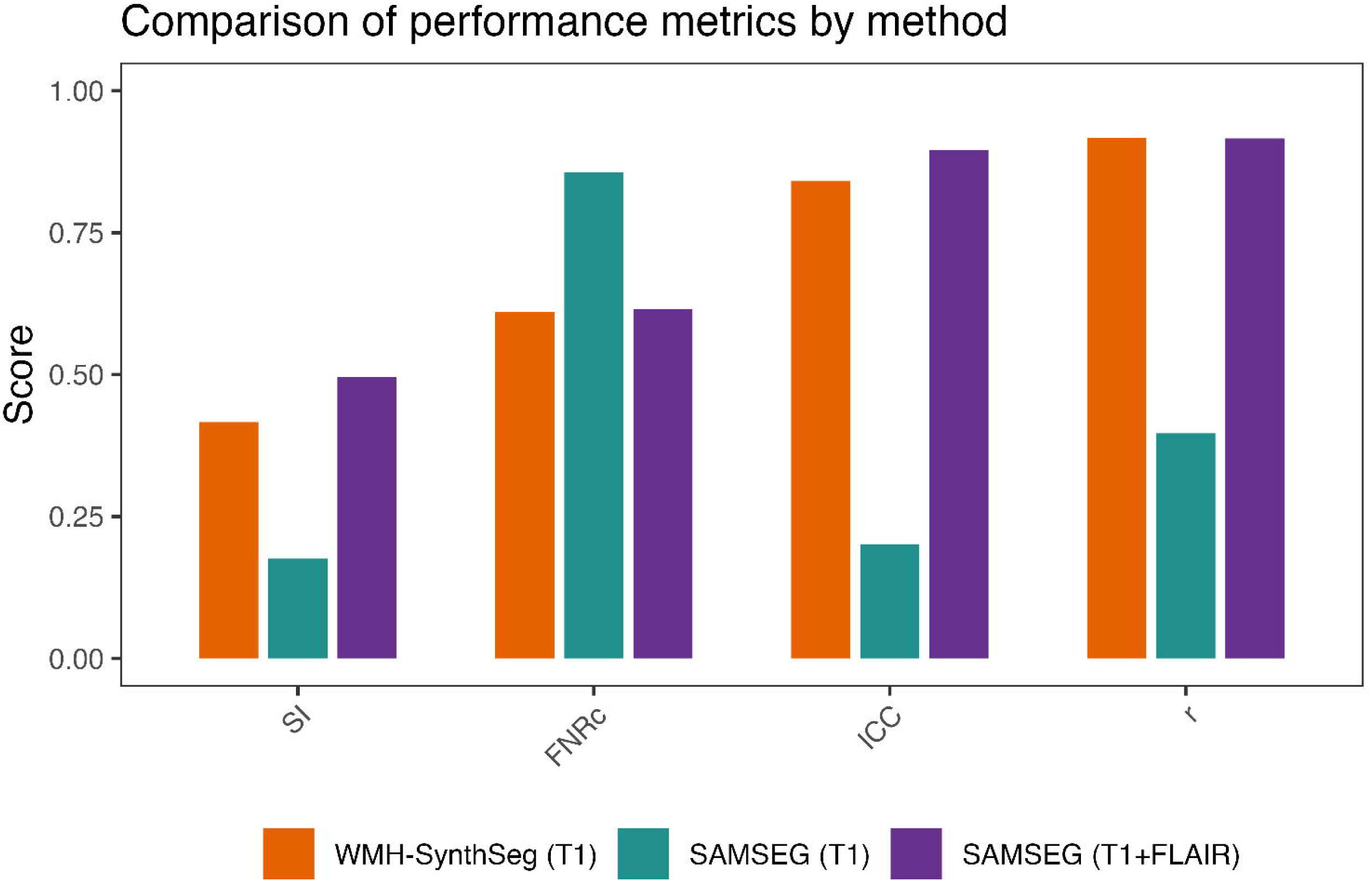
**Comparison of performance metrics for automated WMH masks**. WMH masks generated using WMH-SynthSeg (T1), SAMSEG (T1), and SAMSEG (T1+FLAIR) are compared against the gold standard WMH masks for the entire dataset (n=227). WMH-SynthSeg (T1) across the board performed better than SAMSEG (T1). WMH-SynthSeg (T1) showed a similar performance as SAMSEG (T1+FLAIR) in FNRc, ICC, and Pearson’s correlation, but slightly worse in SI. Abbreviations: SI = Dice similarity index, FNRc = cluster-level false negative ratio, ICC = intraclass correlation, r = Pearson’s correlation coefficient.

**Figure 3:**
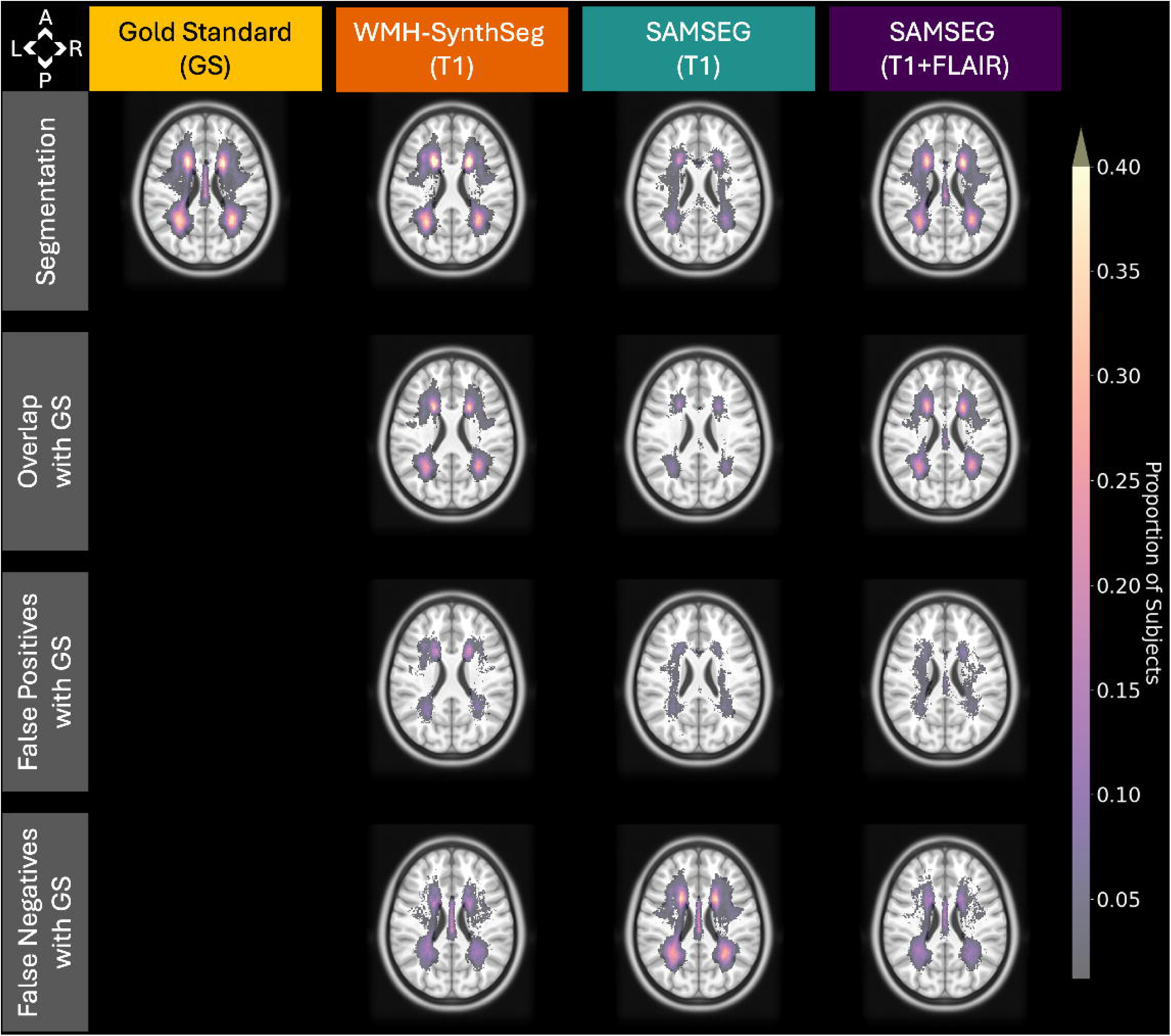
Heatmaps of WMH masks and classification metrics across all subjects, visualized in the MNI152 standard space template (z = +24). All masks were linearly registered to MNI152 space using brain masks as weighting during registration, and resulting alignments were visually inspected for accuracy. The first row shows segmentation heatmaps, the second row shows heatmaps of overlap between WMH-SynthSeg (T1), SAMSEG (T1), and SAMSEG (T1+FLAIR) with the gold standard mask, the third row shows heatmaps of false positives (algorithm incorrectly classified WMH), and the last row shows heatmaps of false negatives (algorithm incorrectly missed WMH). Abbreviations: GS = gold standard, WMH = white matter hyperintensities.

### 3.2. WMH masks generated using WMH-SynthSeg (T1) shows strongest agreement with gold standard WMH volume

A Kruskal-Wallis test revealed a significant difference in volume ratios across the WMH-SynthSeg (T1), SAMSEG (T1), and SAMSEG (T1+FLAIR) segmentation methods (χ²(2) = 127.58, *p* < .001). Pairwise Wilcoxon rank-sum tests showed that all methods differed significantly from one another (*p* < .001 for all comparisons after Bonferroni correction). WMH-SynthSeg (T1) (median = 1.00) had the strongest agreement with the gold standard volumes (Fig. 4). In contrast, SAMSEG (T1) (median = 0.30) and SAMSEG (T1+FLAIR) (median = 0.83) produced systematically lower volume estimates. Effect sizes were large between WMH-SynthSeg (T1) and SAMSEG (T1) (*r* = 0.51), a moderate difference between SAMSEG (T1) and SAMSEG (T1+FLAIR) (r = 0.35), and a small-to-moderate difference between WMH-SynthSeg (T1) and SAMSEG (T1+FLAIR) (r = 0.19).

**Figure 4:**
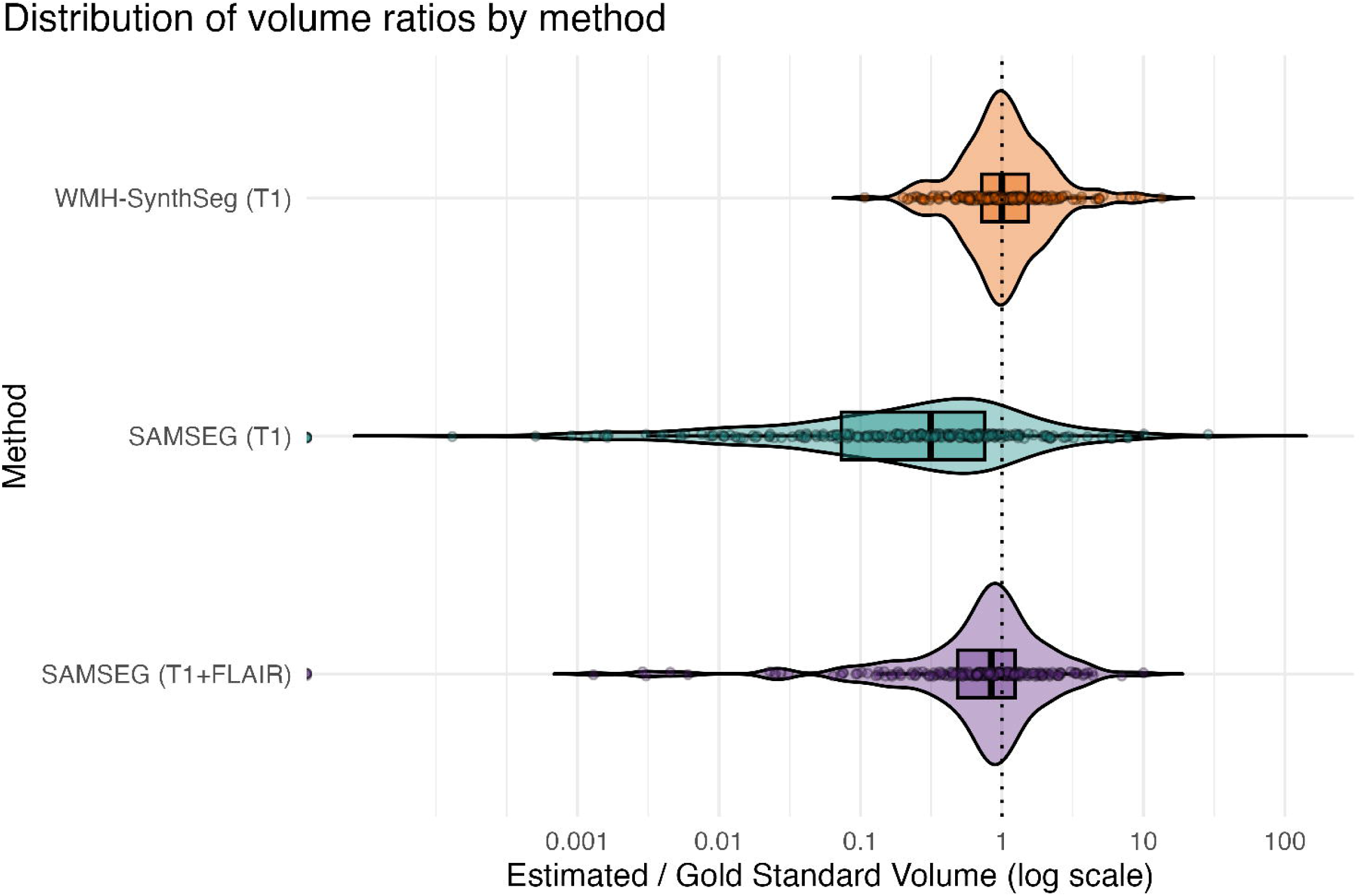
Distribution plots of volume ratio (volume estimated by each algorithm divided by gold standard volume). A volume ratio of 1, noted with a dotted vertical line, indicates perfect agreement. WMH-SynthSeg (T1) demonstrates the highest agreement with the gold standard, while SAMSEG (T1) and SAMSEG (T1+FLAIR) generally underestimate volume. Values are shown on a log-transformed x-axis for interpretability.

### 3.3. WMH-WM contrast levels differ by dataset, independent of age, WMH volume, or stroke lesion volume

WMH-WM intensity ratios, representing the contrast between WMH and normal-appearing WM, varied significantly by dataset (Kruskal–Wallis χ²(2) = 114.68, p < .001), with post hoc Wilcoxon rank-sum tests confirming significant differences between all dataset pairs (p < .001, Bonferroni-corrected). On average, Dataset 1 had the lowest WMH-WM intensity ratio (0.38 ± 0.10), indicating higher contrast between WMH and WM (Fig. 5). In contrast, Dataset 3 exhibited the highest average ratio (0.88 ± 0.23), reflecting both lower and more variable contrast. Dataset 2 showed intermediate values (0.66 ± 0.15). A standardized linear mixed effects regression model (marginal R^2^ = 0.021, conditional R^2^ = 0.692) showed that WMH-WM intensity ratio was not associated with age, sex, days since stroke, WMH volume or stroke lesion volume (Supplementary Table 2).

**Figure 5:**
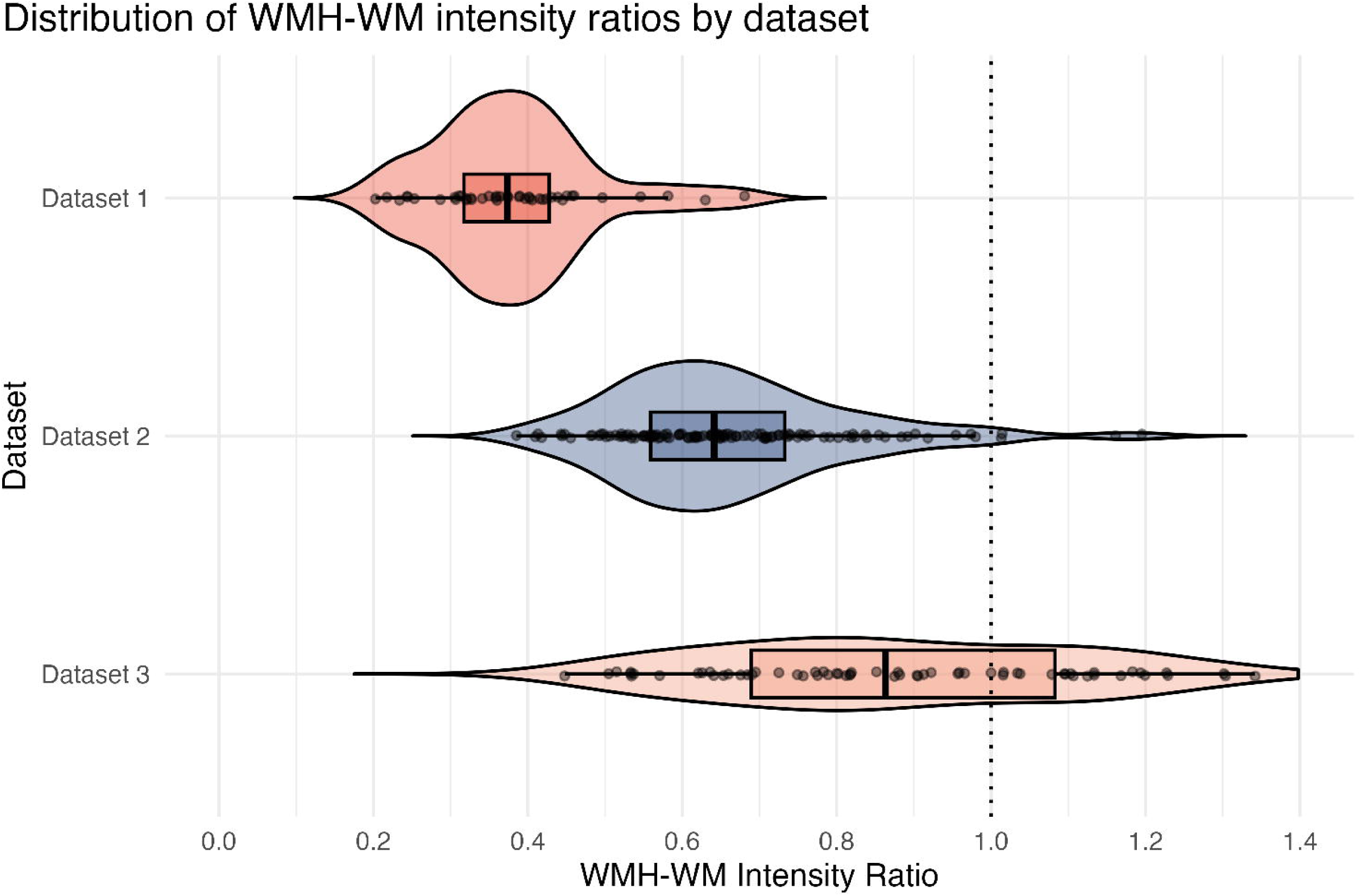
Distribution of WMH-WM intensity ratios for each dataset, where 1 indicates the same intensity for gold-standard WMH and WM voxels. WM voxels were corrected to exclude any voxels that were identified as WMH and stroke lesion. Dataset 1, which consists of individuals with chronic stroke, has the lowest average ratio, and thereby highest average contrast, between WMH and WM tissue. Dataset 3, representing individuals with early subacute stroke, exhibits the highest average ratio and largest range, suggesting poorer and more variable contrast between WMH and WM tissue. Abbreviations: WM = white matter, WMH = white matter hyperintensities.

### 3.4. WMH segmentation accuracy improved with greater WMH volume and WMH-WM contrast but is not related to stroke lesion volume

We used a standardized linear mixed effects regression model to evaluate whether WMH segmentation accuracy, measured by SI, for each algorithm was related to WMH volume, stroke lesion volume, and WMH-WM contrast, adjusting for age, days since stroke, and a random effect of dataset (Fig. 6, Supplementary Table 3). Better WMH segmentation performance was related to higher WMH volumes for all three methods: WMH-SynthSeg (T1) (Β = 0.758, 95% confidence interval [CI]: 0.677 to 0.838, p < 0.001), SAMSEG (T1) (Β = 0.548, 95% CI: 0.450 to 0.646, p < 0.001), and SAMSEG (T1+FLAIR) (Β = 0.659, 95% CI: 0.572 to 0.746, p < 0.001). Better WMH segmentation performance was also related to lower WMH-WM intensity ratios across all three methods: WMH-SynthSeg (T1) (Β = -0.452, 95% CI: -0.553 to -0.351, p < 0.001), SAMSEG (T1) (Β = -0.251, 95% CI: -0.376 to -0.126, p < 0.001), and SAMSEG (T1+FLAIR) (Β = -0.323, 95% CI: -0.432 to -0.214, p < 0.001). In contrast, associations between SI and stroke lesion volume were non-significant for WMH-SynthSeg (T1), SAMSEG (T1), and SAMSEG (T1+FLAIR). Age was a significant covariate for WMH-SynthSeg (T1) (Β = 0.091, 95% CI: 0.012 to 0.171, p = 0.025) and SAMSEG (T1+FLAIR) (Β = 0.258, 95% CI: 0.172 to 0.344, p < 0.001), but not SAMSEG (T1). Sex and days since stroke were non-significant across methods.

**Figure 6:**
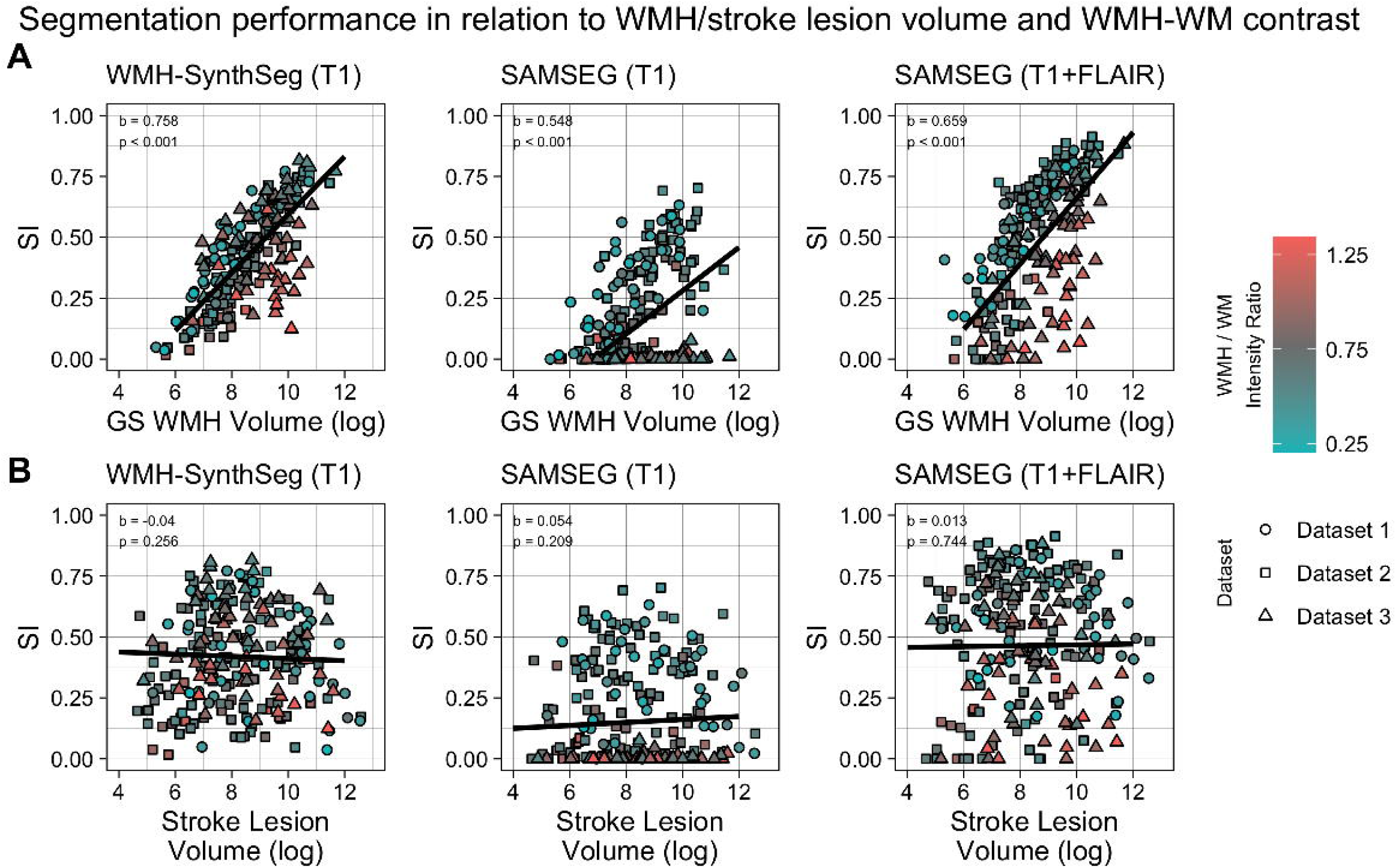
Influence of WMH, stroke lesion volume, and WMH-WM contrast on WMH segmentation performance assessed using Dice similarity index (SI). Linear mixed-effects regression lines, standardized main effect coefficients, and p-values are reported for (A) log-transformed WMH volume and SI, and (B) log-transformed lesion volume and SI. Individual points are colored by WMH-WM intensity ratio, with blue indicating stronger contrast and red indicating weaker contrast. Other covariates in the model were log-transformed WMH-WM intensity ratios, age, sex, log-transformed days since stroke, and a random effect for dataset. Segmentation overlap increases with larger WMH volumes and greater WMH-WM contrast. In contrast, stroke lesion volume shows no significant impact on WMH segmentation quality for WMH-SynthSeg (T1), SAMSEG (T1), and SAMSEG (T1+FLAIR). Abbreviations: GS = gold standard, WM = white matter, WMH = white matter hyperintensities.

## 4. Discussion

In this study, we show that WMH volume in stroke survivors can be reliably estimated using T1 imaging alone, achieving performance comparable to established multimodal methods. Volume estimates from WMH-SynthSeg (T1) showed good consistency and strong agreement with manual tracings, suggesting that T1-only approaches may be a viable alternative for WMH quantification when multimodal imaging is unavailable. However, spatial overlap metrics revealed limitations in accurate WMH localization, indicating that spatial accuracy may benefit from additional imaging modalities or further algorithmic refinement.

Our findings demonstrate that WMH-SynthSeg (T1) substantially outperforms SAMSEG (T1) using the same T1-only input across multiple independent stroke cohorts spanning different recovery stages. WMH-SynthSeg (T1) consistently achieved better classification metrics (SI, FNRc) and greater agreement of volume estimates (ICC, Pearson’s correlation, volume ratio) with the gold standard segmentations compared to SAMSEG (T1). These findings may reflect the differences in model training for each algorithm: the CNN architecture in WMH-SynthSeg benefits from synthetic training explicitly modeling single-modality inputs, enabling robust WMH delineation from T1 images without further retraining (Laso et al., 2024), whereas the generative Bayesian framework in SAMSEG relies heavily on multimodal contrasts to define priors and tissue signatures (Cerri et al., 2021), potentially limiting its accuracy when only T1 data are available.

Performance by WMH-SynthSeg using T1-only input was comparable to SAMSEG using both T1 and FLAIR as input. Across multiple cohorts, WMH-SynthSeg (T1) matched SAMSEG (T1+FLAIR) in segmentation quality based on SI and ICC, even without multimodal information. These findings highlight the ability of WMH-SynthSeg to robustly estimate WMH volume without the need for FLAIR or other complementary sequences. Taken together, our results support the use of WMH-SynthSeg in research settings where multimodal MRI, especially FLAIR, is unavailable, broadening the applicability of automated WMH segmentation in large-scale and retrospective stroke recovery studies.

WMH segmentation performance for both WMH-SynthSeg and SAMSEG increased with higher WMH volumes, consistent with previous work showing that smaller WMH volumes are more challenging for automated algorithms to accurately delineate. This difficulty with smaller WMH volumes has been noted for a range of WMH segmentation tools (Ferris et al., 2023; Kuijf et al., 2019), as small discrepancies can disproportionately impact spatial metrics when WMH volume is limited. Importantly, our results also showed no association between segmentation accuracy and total stroke lesion volume, indicating that even the presence of large stroke lesions did not adversely affect the performance of either algorithm. This robustness underscores the suitability of both WMH-SynthSeg and SAMSEG for WMH quantification in stroke populations with a wide range of lesion volumes.

WMH-SynthSeg also demonstrated robust performance across a wide range of WMH-WM contrast levels, a common source of variability in imaging due to differences in scanners, acquisition protocols, or individual patient characteristics. Our full dataset incorporated images acquired from multiple scanners, each with their own protocols. While contrast varied between datasets, with the early subacute dataset having the widest range, these differences were not explained by age, days since stroke, WMH volume, nor stroke lesion volume. Nevertheless, WMH-SynthSeg maintained acceptable segmentation quality across all datasets. This likely reflects the model’s synthetic training approach, which is designed to enhance the algorithm’s robustness across diverse imaging conditions. One limitation of our analysis, however, is that we only evaluated WMH-SynthSeg using high-resolution T1 images; future work will be necessary to determine whether reliable WMH quantification can be similarly achieved with low-resolution T1 sequences typical of clinical scanners.

Interestingly, WMH-SynthSeg (T1) outperformed WMH-SynthSeg (FLAIR), likely reflecting how the model was trained. Although the ground truth segmentations to guide training were derived from FLAIR, the training data consisted of synthetic T1 images augmented to vary resolution, intensities, orientation, and bias fields. These augmentations make the network largely modality-agnostic, but because synthetic WMHs are generated based on T1 intensity patterns, the model may be effectively learning T1 representations of WMHs rather than true FLAIR signal characteristics. Although WMH volumes from T1 and FLAIR are strongly correlated (Wei et al., 2019), lesions appear differently on each modality. On FLAIR, WMH appear as distinctly bright areas that encompass a wide range of tissue damage severity, whereas on T1, they are visible as dark areas typically in cases of significant injury (Riphagen et al., 2018; Wei et al., 2019). Training on T1 representations likely enhances segmentation accuracy for T1-only data, enabling detection of more subtle WMHs in T1. However, the model may be more liberal in labeling WMH when applied to FLAIR since mild WMH appear differently on T1 than FLAIR, potentially contributing to the overestimation and reduced accuracy we observed on FLAIR-only segmentation. While our results suggest that using T1-only input with WMH-SynthSeg is a viable alternative when multimodal data is unavailable, future work should investigate whether generating synthetic training data from numerous MRI modalities may further improve WMH segmentation and generalizability.

Our study has other limitations despite demonstrating reliable WMH volume estimation using T1-only input. We found that WMH-SynthSeg showed poor to moderate performance in spatial localization. Accurate spatial localization is important because deep and periventricular WMHs have distinct clinical implications and underlying pathologies (Griffanti et al., 2018; Lopes et al., 2025). Improving spatial precision is therefore essential for studies that focus on lesion location-specific effects or on monitoring WMH subtype progression.

We also observed strong collinearity between days since stroke and dataset, as our study included three cohorts representing distinct post-stroke stages (early subacute, late subacute, and chronic). Segmentation performance was related to WMH contrast but showed no direct association with days since stroke, which may be attributable to collinearity with dataset. However, WMH contrast itself differed across datasets, with the chronic cohort demonstrating greater contrast that may have facilitated WMH segmentation. These findings are important considering the profound and ongoing brain reorganization after stroke, including edema, gliosis, and tissue remodeling, which can substantially alter WMH characteristics and their appearance relative to normal-appearing white matter (Riphagen et al., 2018). Future research should incorporate more heterogeneous datasets spanning a wider range of post-stroke stages to better determine whether time since stroke influences WMH segmentation performance.

This work highlights the potential of using WMH-SynthSeg with T1 images only as an effective tool for WMH volume estimation when FLAIR MRI is not available. By leveraging a synthetic training strategy, the algorithm shows robustness across even diverse stroke lesion volumes, addressing common challenges in large-scale studies. Continued improvements in spatial localization and the ability to differentiate stroke lesions from WMHs will be key to advancing fully automated workflows and enhancing clinical utility. This work establishes important groundwork for expanding automated WMH assessment to broader research applications, thereby enabling more detailed investigations of WMH volume as a biomarker for stroke recovery.

## Data Availability

WMH-SynthSeg and SAMSEG are available as part of the open-source FreeSurfer software package. Additional data and code from this study are available upon reasonable request from the corresponding author.

## Data and Code Availability

WMH-SynthSeg and SAMSEG are available as part of the open-source <u>FreeSurfer</u> <u>software package</u>. Additional data and code from this study are available upon reasonable request from the corresponding author.

## Acknowledgements

We would like to acknowledge Dr. Nicolas Schweighofer and Dr. Carolee Winstein for their help with proofreading this manuscript.

## Supplementary Material

Supplementary material is available online.

