## Supplementary Materials for "Estimating white matter hyperintensities volume in individuals with stroke using T1-weighted images"

### Supplementary Material

#### Supplementary Methods

##### Automated WMH segmentation using FLAIR-only inputs

We generated WMH masks using FLAIR-only input to WMH-SynthSeg and SAMSEG, hereby referred to as *WMH-SynthSeg (FLAIR)* and *SAMSEG (FLAIR)* WMH masks (see Supplementary Fig. 1 for a schematic). This allowed us to directly compare WMH-SynthSeg and SAMSEG under identical FLAIR-only conditions and to estimate how much accuracy may be lost when FLAIR is not available. As with the other masks, both sets were corrected to exclude overlapping voxels with the manually traced stroke lesion masks.

#### Supplementary Results

##### WMH-SynthSeg (T1) performs consistently across datasets

WMH-SynthSeg (T1) showed the most consistent performance in individual cohorts, with similar trends observed across individual datasets (Supplementary Fig. 2).

In Dataset 1, both WMH-SynthSeg (T1) (SI = 0.43 ± 0.20, FNRc = 0.40 ± 0.16, ICC = 0.95, r = 0.98) and SAMSEG (T1+FLAIR) (SI = 0.53 ± 0.18, FNRc = 0.33 ± 0.21, ICC = 0.85, r = 0.92) achieved moderate-to-good accuracy and agreement with the gold standard segmentations, while SAMSEG (T1) showed reduced accuracy and agreement (SI = 0.28 ± 0.19, FNRc = 0.75 ± 0.16, ICC = 0.65, r = 0.76).

Similar trends were observed in Dataset 2 (n = 120), where WMH-SynthSeg (T1) (SI = 0.38 ± 0.20, FNRc = 0.73 ± 0.10, ICC = 0.94, r = 0.98) achieved moderate-to-good accuracy and agreement, outperforming SAMSEG (T1) (SI = 0.23 ± 0.21, FNRc = 0.88 ± 0.14, ICC = 0.51, r = 0.79) and performing similarly to SAMSEG (T1+FLAIR) (SI = 0.53 ± 0.28, FNRc = 0.73 ± 0.19, ICC = 0.97, r = 0.97).

In Dataset 3 (n = 64), WMH-SynthSeg (T1) maintained robust performance (SI = 0.48 ± 0.18, FNRc = 0.53 ± 0.18, ICC = 0.72, r = 0.93), often outperforming SAMSEG (T1+FLAIR) (SI = 0.42 ± 0.25, FNRc = 0.58 ± 0.20, ICC = 0.83, r = 0.91). Meanwhile, performance of SAMSEG (T1) markedly deteriorated across all metrics (SI = 0.004 ± 0.001, FNRc = 0.89 ± 0.24, ICC ≈ 0, r = -0.09).

##### Performance of WMH masks produced using FLAIR-only inputs

We evaluated the performance of WMH-SynthSeg and SAMSEG using FLAIR-only input (Supplementary Figure 3). WMH-SynthSeg (FLAIR) achieved poor performance across all datasets (SI = 0.36 ± 0.23, FNRc = 0.49 ± 0.16, ICC = 0.49, r = 0.57). SAMSEG (FLAIR) achieved moderate performance in relation to the gold standard across all datasets (SI = 0.45 ± 0.28, FNRc = 0.67 ± 0.29, ICC = 0.64, r = 0.69).

We performed a Kruskal–Wallis test using the volume ratios from all five automated methods, finding significant group differences (χ²(4) = 319.19, p < .001). Pairwise Wilcoxon rank-sum tests indicated that most methods differed significantly from one another (all p < .001 after Bonferroni correction), except for SAMSEG (FLAIR) and SAMSEG (T1+FLAIR), which did not differ significantly. WMH-SynthSeg (FLAIR) (median = 2.64) produced systematically higher volume estimates, while SAMSEG (FLAIR) (median = 0.74) demonstrated lower volume estimates. Effect sizes were largest between WMH-SynthSeg (FLAIR) and each of SAMSEG (T1) (r = 0.67), SAMSEG (FLAIR) (r = 0.56) and SAMSEG (T1+FLAIR) (r = 0.53), moderate between WMH-SynthSeg (T1) and WMH-SynthSeg (FLAIR) (r = 0.43), and small to moderate for other pairwise comparisons (r = 0.19–0.35).

#### Supplementary Figures

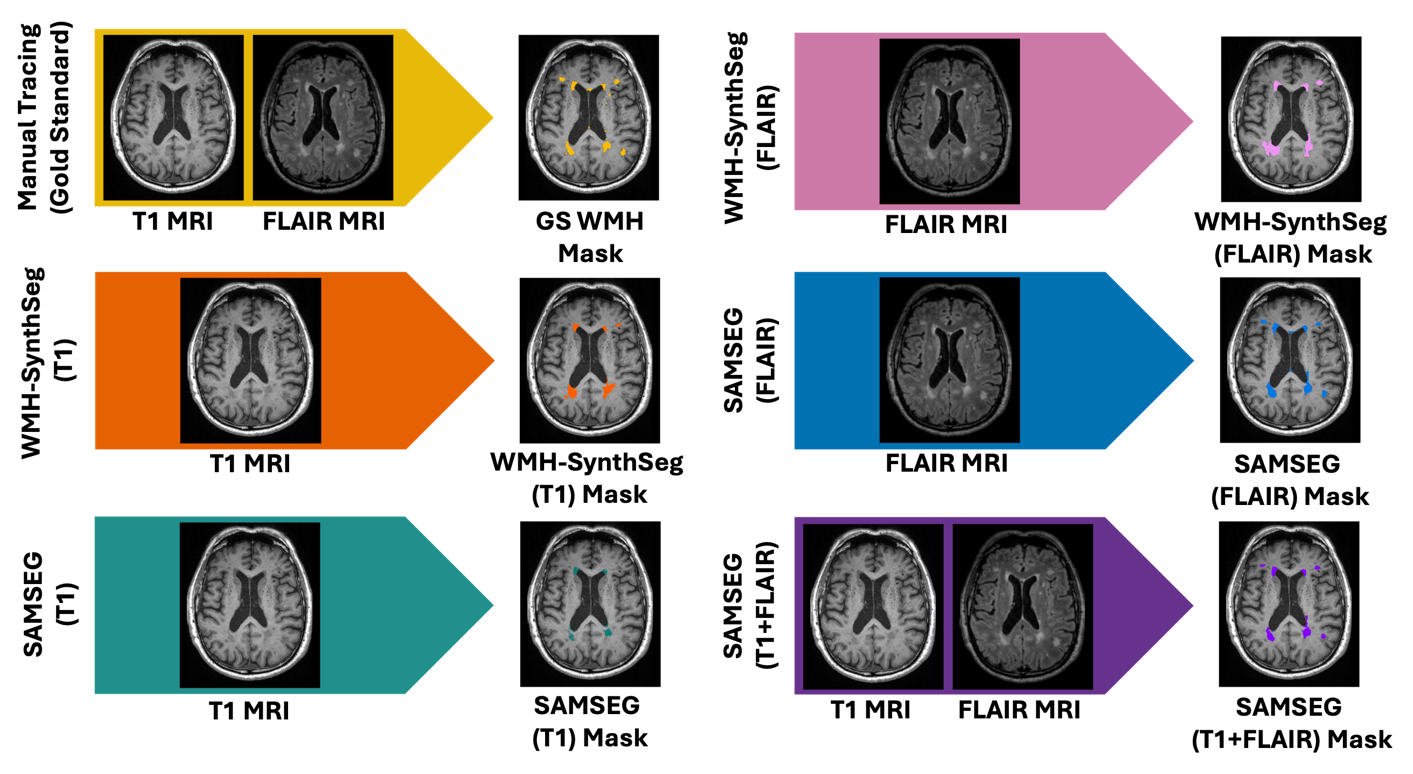

**Supplementary Figure 1: Schematic demonstrating the algorithm and input combinations used to produce WMH masks.** This figure expands on Figure 1 by including two additional methods: WMH-SynthSeg (FLAIR) and SAMSEG (FLAIR). WMH-SynthSeg (FLAIR) and SAMSEG (FLAIR) masks were produced using FLAIR-only input to WMH-SynthSeg and to SAMSEG, respectively.

| Dataset | Scanner Manufacturer | Scanner Model | Sequence | Resolution | TR (ms) | TE (Ms) | Flip Angle | Acquisition Matrix |
| --- | --- | --- | --- | --- | --- | --- | --- | --- |
| Dataset 1 | Phillips | Achieva / Elition | MPRAGE | 1 mm isotropic | 3000 | 3.7 | 90° | 256 x 224 |
|  |  |  | FLAIR | 0.94 x 0.94 x 3 | 9000 | 90 | 90° | 240 x 191 |
| Dataset 2 | Siemens | Tim Trio | MPRAGE | 1 mm isotropic | 1900 | 2.6 | 9° | 256 x 256 |
|  |  |  | FLAIR | 0.5 x 0.5 x 1 | 6000 | 380 | 120° | 512 x 512 |
| Dataset 3 | Siemens | MAGNATOM Verio / MAGNATOM PrismaFit | MPRAGE | 1 mm isotropic | 2300 | 2 / 2.98 / 2.9 | 9° | 224 x 224 / 256 x 240 / 256 x 256 |
|  |  |  | FLAIR | 0.72 x 0.72 x 5 /  1 x 1.2 x 1 /  1 mm isotropic | 5850 / 4800 / 6000 | 83 / 441 / 325 | 150° / 120° | 240 x 320 / 256 x 256 |

**Supplementary Table 1: Imaging parameters for each dataset.** Abbreviations: MPRAGE = magnetization-prepared rapid gradient-echo, FLAIR = fluid-attenuated inversion recovery, TR = repetition time, TE = echo time.

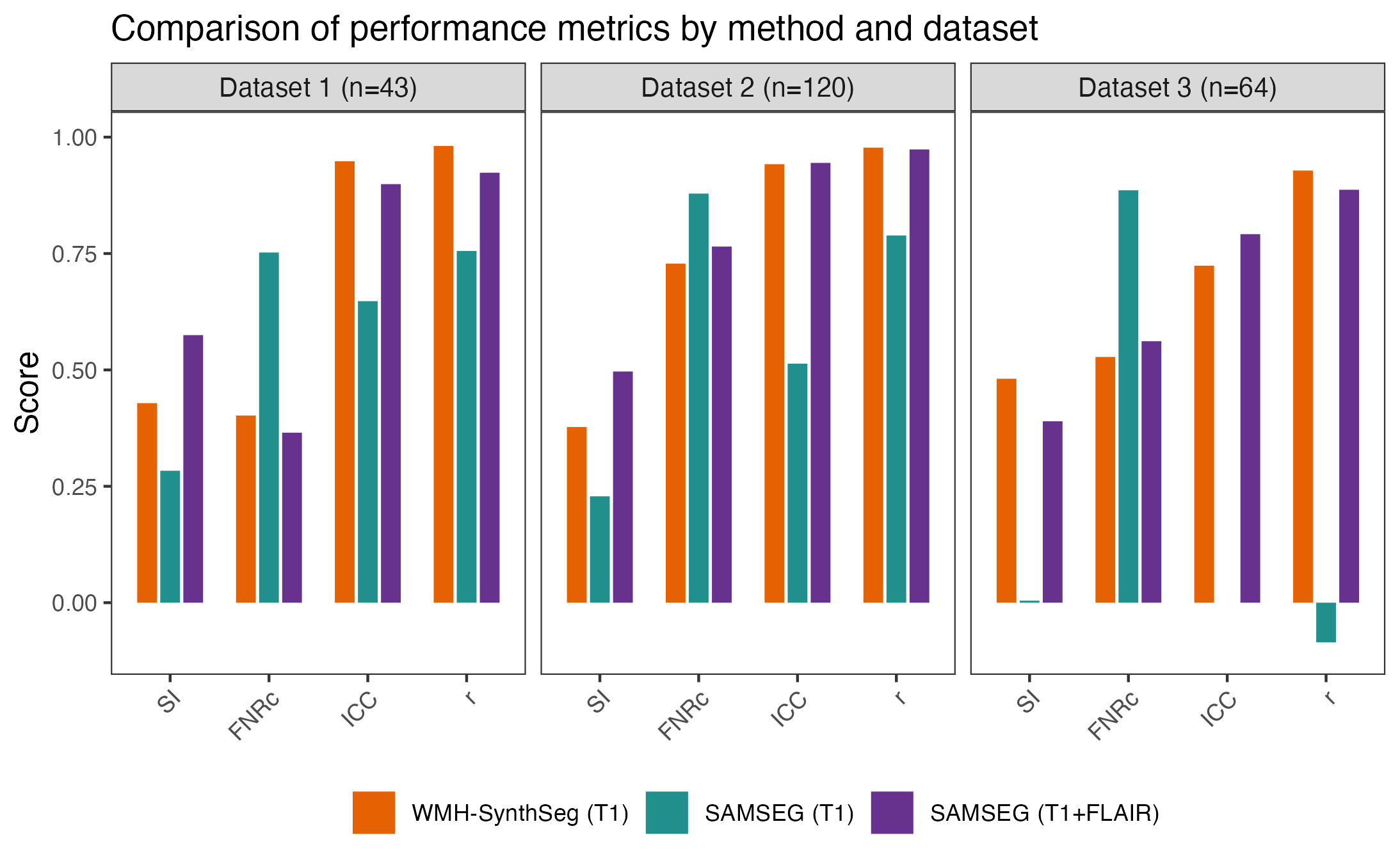

**Supplementary Figure 2: Comparison of performance metrics by method and dataset.** Masks generated using WMH-SynthSeg (T1), SAMSEG (T1), and SAMSEG (T1+FLAIR) are compared against the gold standard WMH masks for each dataset. In each dataset, WMH-SynthSeg (T1) compared more favorably to the gold standard than SAMSEG (T1) did across all metrics. WMH-SynthSeg (T1) also performed comparably to SAMSEG (T1+FLAIR). In Dataset 1, WMH-SynthSeg (T1) had better consistency metrics (ICC, Pearson correlation) but slightly worse classification metrics (SI, FNRc) to SAMSEG (T1+FLAIR). In Dataset 2, WMH-SynthSeg (T1) showed improved performance over SAMSEG (T1+FLAIR) in FNRc and Pearson correlation, matched SAMSEG (T1+FLAIR) in ICC, but had lower SI scores. In Dataset 3, WMH-SynthSeg (T1) outperformed SAMSEG (T1+FLAIR) in SI, FNRc, and Pearson correlation, while exhibiting a slightly lower ICC. Abbreviations: SI = Dice similarity index, FNRc = cluster-level false negative ratio, ICC = intraclass correlation coefficient, r = Pearson’s correlation coefficient.

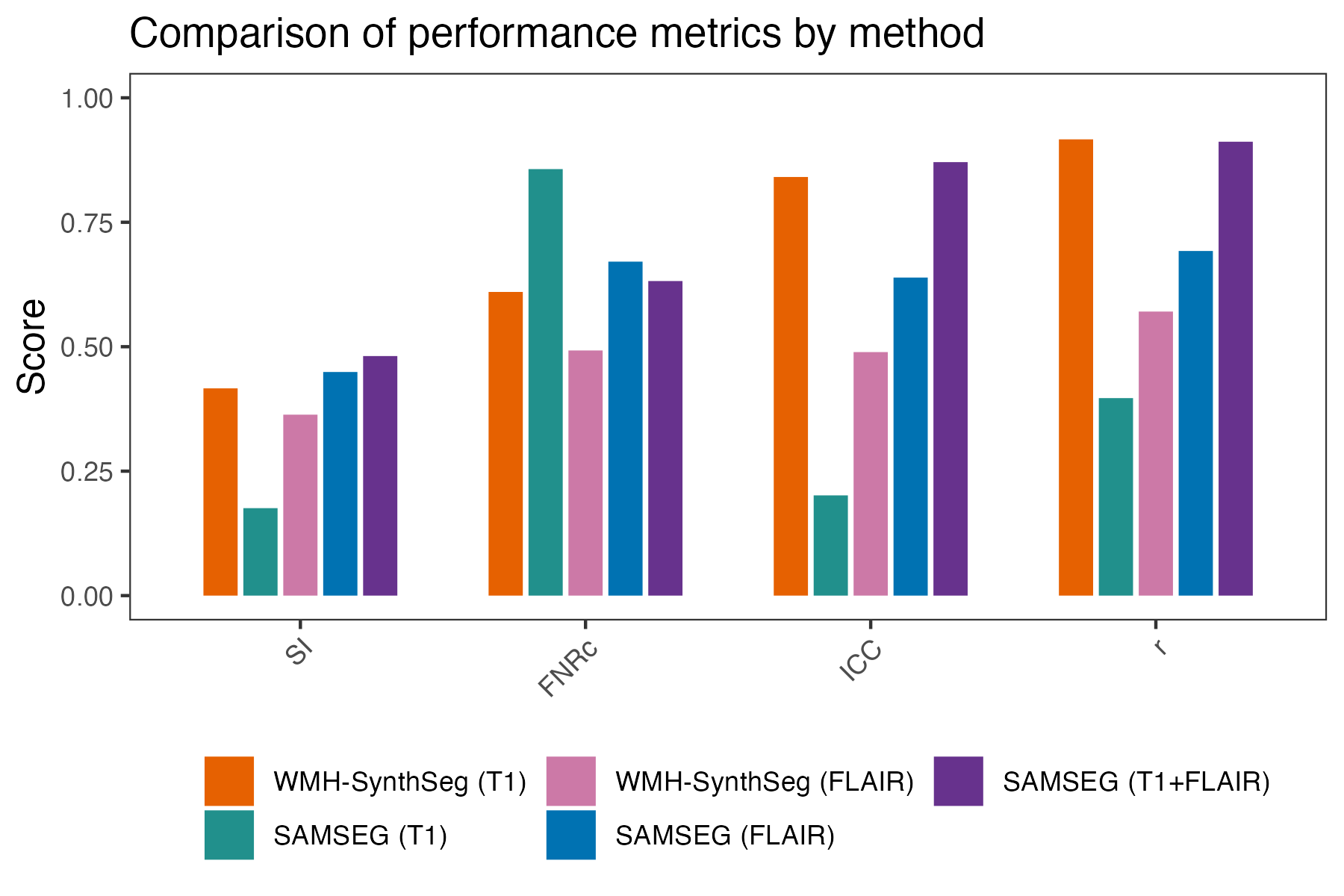

**Supplementary Figure 3: Comparison of performance metrics for automated WMH masks**. WMH masks generated using WMH-SynthSeg (FLAIR) and SAMSEG (FLAIR) are compared against the gold standard WMH masks for the entire dataset (n = 227). This figure expands on Figure 2 by including results from all five methods: WMH-SynthSeg (T1), SAMSEG (T1), WMH-SynthSeg (FLAIR), SAMSEG (FLAIR), and SAMSEG (T1+FLAIR). SAMSEG (FLAIR) performed better than WMH-SynthSeg (FLAIR) in all metrics except FNRc. WMH-SynthSeg (T1) outperformed both WMH-SynthSeg (FLAIR) and SAMSEG (FLAIR) across all metrics, except for FNRc for WMH-SynthSeg (FLAIR) and SI for SAMSEG (FLAIR). Abbreviations: SI = Dice similarity index, FNRc = cluster-level false negative ratio, ICC = intraclass correlation, r = Pearson’s correlation coefficient.

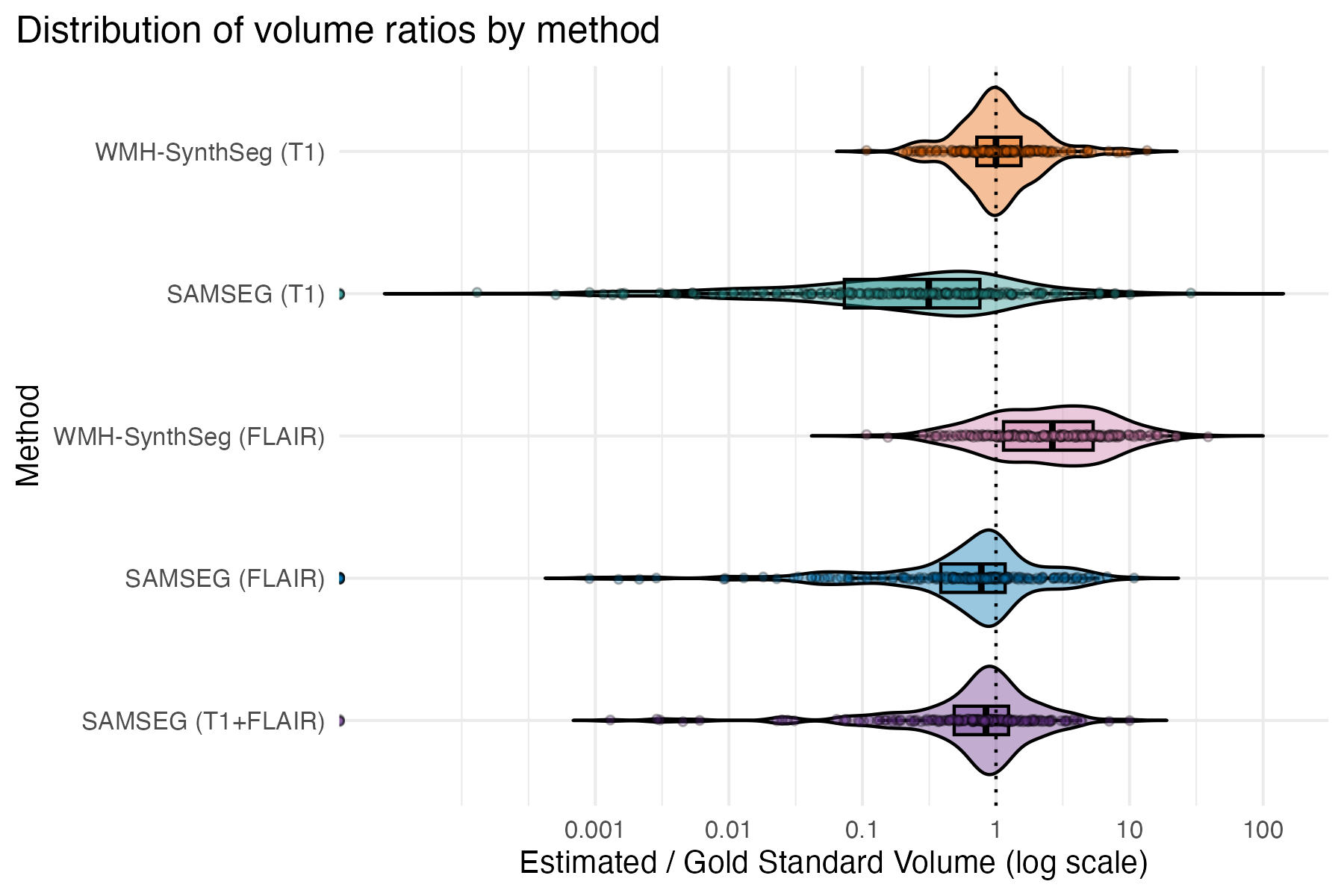

**Supplementary Figure 4: Expanded distribution plots of volume ratio (volume estimated by each algorithm divided by gold standard volume).** This figure expands on Figure 4 by including results from all five methods: WMH-SynthSeg (T1), SAMSEG (T1), WMH-SynthSeg (FLAIR), SAMSEG (FLAIR), and SAMSEG (T1+FLAIR). A volume ratio of 1, noted with a dotted vertical line, indicates perfect agreement. WMH-SynthSeg (T1) demonstrates the highest agreement with the gold standard. WMH-SynthSeg (FLAIR) tends to overestimate the volume. SAMSEG (FLAIR), like SAMSEG (T1) and SAMSEG (T1+FLAIR), generally underestimates volume. Values are shown on a log-transformed x-axis for interpretability.

| WMH-WM Intensity Ratio  N = 227, Marginal R^2^ = 0.021, Conditional R^2^ = 0.692 | | | | |
| --- | --- | --- | --- | --- |
| Predictors | *B* | SE | 95% CI | *p* |
| Age | -0.090 | 0.052 | -0.192 to 0.012 | 0.083 |
| Sex [Male] | 0.073 | 0.092 | -0.109 to 0.254 | 0.430 |
| log(Days since stroke) | -0.137 | 0.135 | -0.403 to 0.128 | 0.309 |
| log(WMH volume) | -0.035 | 0.053 | -0.139 to 0.069 | 0.504 |
| log(Lesion volume) | 0.033 | 0.046 | -0.057 to 0.123 | 0.464 |

**Supplementary Table 2: Summary of standardized linear mixed effects regression model predicting WMH-WM intensity ratio (n = 227, marginal R² = 0.021).** Contrast between WMH and normal-appearing WM did not have any significant associations with age, sex, days since stroke, gold standard WMH volume, or stroke lesion volume. Abbreviations: WM = white matter, WMH = white matter hyperintensities.

| Dice Similarity Index with gold standard WMH mask  N = 227 | | | | | | | | | | | | |
| --- | --- | --- | --- | --- | --- | --- | --- | --- | --- | --- | --- | --- |
|  | WMH-SynthSeg (T1)  Marginal R^2^ = 0.697  Conditional R^2^ = 0.747 | | | | SAMSEG (T1) Marginal R^2^ = 0.239  Conditional R^2^ = 0.742 | | | | SAMSEG (T1+FLAIR) Marginal R^2^ = 0.661  Conditional R^2^ = 0.708 | | | |
| Predictors | *B* | SE | 95% CI | *p* | *B* | SE | 95% CI | *p* | *B* | SE | 95% CI | *p* |
| log(WMH-WM intensity ratio) | -0.452 | 0.051 | -0.553  to  -0.351 | **<0.001** | -0.251 | 0.063 | -0.376  to  -0.126 | **<0.001** | -0.323 | 0.055 | -0.432  to  -0.214 | **<0.001** |
| log(GS WMH volume) | 0.758 | 0.041 | 0.677  to  0.838 | **<0.001** | 0.548 | 0.050 | 0.450  to  0.646 | **<0.001** | 0.659 | 0.044 | 0.572  to  0.746 | **<0.001** |
| log(Stroke lesion volume) | -0.040 | 0.036 | -0.110  to  0.030 | 0.256 | 0.054 | 0.043 | -0.031  to  0.139 | 0.209 | 0.013 | 0.039 | -0.063  to  0.089 | 0.744 |
| Age | 0.091 | 0.040 | 0.012  to  0.171 | **0.025** | 0.046 | 0.049 | -0.051  to  0.143 | 0.351 | 0.258 | 0.044 | 0.172  to  0.344 | **<0.001** |
| Sex (Male) | 0.009 | 0.072 | -0.133  to  0.150 | 0.902 | 0.132 | 0.087 | -0.039  to  0.304 | 0.129 | 0.099 | 0.078 | -0.054  to  0.253 | 0.204 |
| log(Days since stroke) | -0.064 | 0.085 | -0.230  to  0.103 | 0.453 | -0.061 | 0.127 | -0.310  to  0.189 | 0.632 | 0.074 | 0.088 | -0.098  to  0.246 | 0.399 |

**Supplementary Table 3:** **Summary of standardized linear mixed-effects regression models predicting the Dice similarity index with respect to the gold standard WMH masks for segmentations produced by WMH-SynthSeg (T1), SAMSEG (T1), and SAMSEG (T1+FLAIR).** Fixed effects included the log-transformed WMH-WM intensity ratio, log-transformed gold standard WMH volume, log-transformed stroke lesion volume, age, and log-transformed days since stroke; dataset was modeled as a random effect. WMH-SynthSeg (T1) showed the highest marginal R² (0.697), indicating that the fixed effects explained more variance in segmentation accuracy compared to SAMSEG (T1) (marginal R² = 0.239) and SAMSEG (T1+FLAIR) (marginal R² = 0.661). Higher SI was associated with lower WMH-WM intensity ratios for all three methods. As observed in Figure 4, SI was significantly associated with WMH volume. SI was associated with age for WMH-SynthSeg (T1) and SAMSEG (T1+FLAIR), while sex, lesion volume and days since stroke were not significant predictors for any algorithm. Abbreviations: GS = gold standard, WM = white matter, WMH = white matter hyperintensities.
